# Neurological consequences of ischemic and hemorrhagic stroke in the UK Biobank

**DOI:** 10.64898/2026.01.25.26344805

**Authors:** Ambika Grover, David A. Bennett, Venkatesh L. Murthy, Chirag J. Patel

## Abstract

**Introduction:** Stroke is associated with long-term neurological complications, including mortality, cognitive impairment, dementia, and structural brain injury, but these outcomes are typically studied in isolation. We evaluated multiple dimensions of post-stroke neurological burden within a large population-based cohort.

**Methods:** Using UK Biobank data, we evaluated associations between incident stroke and long-term outcomes across survival, dementia, cognitive performance, and white matter hyperintensity (WMH) burden. Time-to-event outcomes were assessed using multivariable Cox proportional hazards models and Fine–Gray competing-risk models for dementia. Cognitive and neuroimaging outcomes were evaluated using covariate-adjusted linear models.

**Results:** Among 14,806 participants with incident stroke (69.2% ischemic, 11.5% intracerebral hemorrhage [ICH], 9.8% other nontraumatic intracranial hemorrhage [ONIH], and 9.5% subarachnoid hemorrhage [SAH]), stroke was associated with substantially elevated mortality (hazard ratios ranging from 3.89 for SAH to 7.62 for ICH versus stroke-free person-time). Dementia risk was increased across stroke subtypes; however, cumulative dementia incidence remained modest relative to mortality, reflecting substantial competing mortality. Cognitive differences were observed years after stroke, with slower reaction times observed broadly and lower reasoning performance in subsets of stroke survivors. WMH burden was higher following stroke, particularly among individuals with ischemic stroke and ICH. Although outcome severity varied across individuals and stroke types, substantial overlap was observed across survival, cognitive, neurodegenerative, and neuroimaging domains.

**Conclusions:** Neurological consequences following stroke extend across multiple dimensions of brain health and cannot be fully captured by any single outcome measure. Integrating survival, cognition, dementia, and neuroimaging phenotypes provides a more comprehensive characterization of post-stroke prognosis.

## Introduction

Stroke is often conceptualized as an acute neurological event, yet its consequences unfold over years and span multiple domains of brain health.^1,2^ Stroke is well known to constitute a significant global burden.^3^ Although mortality, dementia, cognitive impairment, and structural brain injury have each been linked to stroke, these outcomes are typically investigated separately and rarely evaluated within the same population.^4–11^ Consequently, the overall neurological burden of stroke remains incompletely characterized, and it is unclear how different manifestations of post-stroke neurological injury relate to one another over time.

Among nontraumatic stroke types—including acute ischemic stroke (AIS), subarachnoid hemorrhage (SAH), intracerebral hemorrhage (ICH), and other nontraumatic intracranial hemorrhage (ONIH)—long-term outcomes have been examined across clinical, cognitive, and neuroimaging domains.^12–14^ However, mortality, dementia, cognitive performance, and neuroimaging markers are rarely evaluated together, despite representing related dimensions of post-stroke neurological health. As a result, it remains difficult to place individual outcomes into a broader context or to understand how different manifestations of post-stroke neurological burden coexist among survivors.

Leveraging linked clinical, cognitive, and neuroimaging data from more than 14,000 UK Biobank participants with incident nontraumatic stroke,^15,16^ we examined long-term neurological outcomes across survival, dementia, cognitive performance, and white matter hyperintensity burden. Rather than evaluating these consequences in isolation, we integrated them within a single population-based cohort to characterize the long-term neurological burden of stroke across multiple dimensions of brain health.

## Methods

### Study Population and Inclusion Criteria

The UK Biobank (UKB) is a population-based cohort of over 500,000 adults recruited from England, Scotland, and Wales, with linked genetic, lifestyle, clinical, and biospecimen data.^15,16^ Longitudinal follow-up is supported by repeat assessment-center visits and linkage to hospital and death records. An overview of cohort construction, time origins, and analytic workflows is shown in Figure 1.

**Figure 1.**
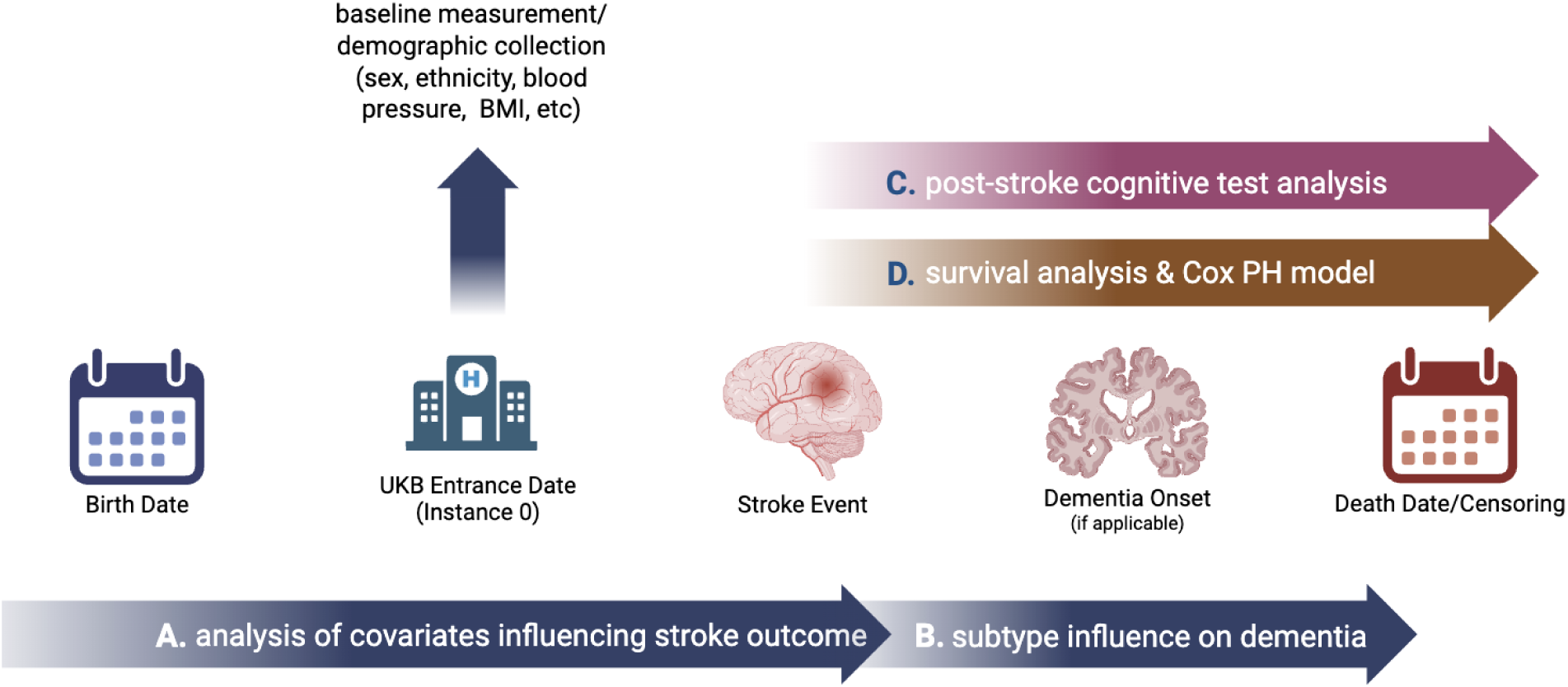
Overview inclusion-exclusion criteria for UKB analysis. (A) Baseline cohort entry at the UK Biobank assessment (instance 0) and ascertainment of demographic, cardiovascular, and genetic covariates (blue/gray arrow). (B) Identification of incident nontraumatic stroke events and classification by subtype—acute ischemic stroke (AIS), intracerebral hemorrhage (ICH), subarachnoid hemorrhage (SAH), and other nontraumatic intracranial hemorrhage (ONIH)—based on linked hospital records (light gray arrow). (C) Cross-sectional post-stroke cognitive assessments, including reaction time and fluid intelligence, among participants with available testing data (purple arrow). (D) Post-stroke time-to-event analyses for all-cause mortality and incident all-cause dementia, including post-stroke survival curves and Cox proportional hazards models with appropriate time origins and censoring (brown arrow).

Among the full UKB cohort (n = 502,244), 15,600 participants experienced at least one nontraumatic stroke. Ischemic stroke was the most common subtype (n = 11,117; 71.3%; mean age at stroke 68.3 years; 39.6% female), followed by intracerebral hemorrhage (ICH; n = 2,633; 16.9%; mean age 67.9 years; 44.9% female), subarachnoid hemorrhage (SAH; n = 2,117; 13.6%; mean age 58.6 years; 61.9% female), and other nontraumatic intracranial hemorrhage (ONIH; n = 1,564; 10.0%; mean age 66.0 years; 37.2% female). Percentages exceed 100% due to individuals experiencing multiple stroke subtypes.

After excluding participants with missing key covariates (age, sex, ethnicity, blood pressure, and body mass index), the analytic stroke subcohort comprised 14,806 individuals. The distribution of stroke subtypes and baseline characteristics was similar between included and excluded participants (Supplementary Table 8). Within the analytic cohort, ischemic stroke accounted for 69.0% of cases (n = 10,251), followed by ICH (11.5%; n = 1,699), ONIH (9.8%; n = 1,448), and SAH (9.5%; n = 1,408).

Stroke events were identified using International Classification of Diseases, Tenth Revision (ICD-10) codes: SAH (I60), ICH (I61), ONIH (I62), and ischemic stroke (I63). All strokes were restricted to nontraumatic etiologies. Records were derived from linked administrative data (hospital and death records) and self-report.^15, 16^ Because historical stroke events prior to data linkage may be incompletely captured, the index stroke was defined as the first recorded stroke within UKB rather than a confirmed first-ever event. Prior work suggests that ICD-10 codes demonstrate moderate sensitivity and high specificity for stroke identification in administrative datasets.^17,18^ ONIH represents a clinically heterogeneous category of nontraumatic intracranial bleeding distinct from ICH and SAH.

To evaluate post-stroke neurological outcomes, we constructed a comparison cohort including both stroke-exposed individuals and stroke-free participants. Individuals without stroke were defined as those without any ICD-10 stroke codes during follow-up. For stroke-exposed participants, follow-up began at the date of the first recorded stroke; for stroke-free participants, follow-up began at the baseline UKB assessment. Differences in follow-up initiation were addressed through age adjustment and time-to-event modeling. This cohort was used to examine associations between stroke and (a) all-cause mortality, (b) incident dementia with mortality as a competing risk, (c) white matter hyperintensity burden, and (d) cognitive outcomes. Neurological outcomes were assessed following the index stroke event, with timing detailed in subsequent sections. Analyses of WMH and cognitive outcomes were restricted to participants with available data, which may preferentially reflect healthier survivors. Baseline covariates were derived from the initial UKB assessment (instance 0) or the closest pre-stroke assessment where applicable.

Although the primary analytic cohort included 14,806 individuals, survival analyses required valid index dates and non-negative follow-up intervals (i.e., death or censoring occurring after the index stroke). Participants with missing or inconsistent time-to-event data were excluded from survival curve analyses, resulting in a reduced sample for these analyses. This reduction reflects data completeness rather than censoring.

### Evaluating post-stroke incidence of dementia

We evaluated the risk of incident dementia, accounting for demographic and cardiovascular risk factors. All-cause dementia was defined using ICD-10 codes F00–F03 and G30, capturing major dementia subtypes including Alzheimer’s disease, vascular dementia, and related disorders. Covariates included age at follow-up start, sex, ethnicity, baseline systolic and diastolic blood pressure, and baseline body mass index (BMI; instance 0). Baseline covariates were used to avoid conditioning on post-stroke physiological changes that could bias estimates of dementia risk. Covariates were selected a priori based on established associations with stroke and neurodegenerative outcomes and their availability within UK Biobank.^6,19^

To account for the temporal sequence of stroke and dementia, we defined each participant’s follow-up start time based on their stroke history. For participants with a history of stroke, follow-up began on the date of their first recorded stroke. For those without stroke, follow-up began at the date of their baseline UK Biobank assessment. Only dementia diagnoses occurring after the fixed date (stroke or baseline) were included in the analysis.

The outcome was time to dementia onset, with participants censored at the earliest of death or the end of follow-up (December 30, 2022). We fit multivariable Cox proportional hazards models comparing stroke-exposed participants to stroke-free participants, with stratified analyses by stroke characteristics, including each stroke subtype, and adjusting for age at the start of follow-up, sex, ethnicity, baseline systolic and diastolic blood pressure, and baseline BMI. Hazard ratios, therefore, reflect post-stroke dementia risk among individuals who survived stroke, relative to stroke-free participants. Formal assessment of proportional hazards using Schoenfeld residuals indicated minor departures from proportionality, as anticipated given the large sample size; the Cox model was therefore retained for primary inference. To complement these population-level estimates, we additionally performed competing risks analyses to estimate dementia incidence among stroke survivors, accounting for mortality.

### Competing risks of post-stroke dementia

Given substantial post-stroke mortality, we performed competing risk analyses treating death prior to dementia as a competing event, as standard survival models may otherwise overestimate dementia incidence.^20^ The analytic cohort included participants with a documented index stroke, defined as the earliest recorded nontraumatic stroke event. Follow-up began at the index stroke and continued until dementia diagnosis, death, loss to follow-up, or administrative censoring. Incident all-cause dementia was defined using ICD-10 codes F00–F03 and G30, and only diagnoses occurring after stroke were included. Participants who remained alive and dementia-free at the end of follow-up were censored. Cumulative incidence functions (CIFs) for dementia and death were estimated nonparametrically and compared across groups using Gray’s test. Absolute risks were reported at 1, 3, 5, 10, and 15 years following stroke.

We fit Fine–Gray subdistribution hazards models to estimate the association between covariates and cumulative incidence of dementia, accounting for competing mortality. The primary model included age at stroke, sex, and ethnicity as core covariates; stroke subtype was included as an exposure variable. In sensitivity analyses, we additionally adjusted for baseline systolic and diastolic blood pressure and body mass index.

As a complementary analysis, we fit cause-specific Cox models treating death as a censoring event to facilitate comparison with standard time-to-event approaches.

### Associations between stroke and post-stroke cognitive performance

Cognitive testing in UK Biobank comprises task-based assessments capturing performance across domains such as processing speed, executive function, memory, and visuospatial reasoning. All tests are administered across various sites using a touchscreen monitor (Supplementary Table 1) or other automated interface (e.g., pressing buttons). For example, the reaction time test assesses an individual’s reaction speed based on 12 rounds of a card game ‘Snap,’ wherein a patient is shown two cards at a time and, if identical, is instructed to press a button box on the table in front of them as quickly as possible. Meanwhile, the tower rearranging task—modeled after the Towers of Hanoi—presents participants with an illustration of three towers containing three differently colored hoops; participants are asked to indicate how many moves it would take to rearrange the hoops into a given configuration.^21,22^

We examined differences in reaction time, numeric memory, reasoning, trail-making, matrix pattern completion, symbol digit substitution, tower rearranging, paired-associate learning, prospective memory, and pairs matching among individuals with different stroke types. Supplementary Table 1 summarizes the function of each of these tests and indicates which output variable from the test was utilized.^21,22^

To evaluate post-stroke cognitive performance among stroke survivors, we excluded participants with a diagnosis of all-cause dementia or parkinsonism before cognitive assessment. For participants with a history of stroke, we identified the closest available cognitive test completed after the index stroke event; stroke-free participants contributed cognitive assessments from available follow-up visits. Cognitive outcomes were analyzed as cross-sectional measures of post-stroke cognitive performance, and the timing of cognitive assessment relative to stroke varied across participants. Stroke subtype was defined based on the most recent recorded stroke event occurring prior to the cognitive assessment. Participants were grouped in two ways: (1) by stroke subtype (ischemic stroke, intracerebral hemorrhage, subarachnoid hemorrhage, or other nontraumatic intracranial hemorrhage) versus no stroke, and (2) by stroke versus no stroke status.

To account for potential confounding, we estimated covariate-adjusted cognitive test scores using linear regression models including age at cognitive assessment (modeled with a quadratic term), sex, ethnicity, and index of deprivation. Adjusted scores were calculated from model residuals and represent cognitive performance after accounting for these covariates. The coefficient of determination (R²) from each model was used to quantify the proportion of variance in cognitive test performance explained by the adjustment variables.

Group differences in adjusted cognitive scores were evaluated using analysis of variance (ANOVA), followed by Tukey post hoc tests to identify pairwise differences between stroke subtype groups and stroke-free participants. Results were summarized separately for each cognitive test.

### Differences in brain image-derived phenotypes across stroke subtypes

Image-derived phenotypes (IDPs) are quantitative measures of brain structure derived from standardized processing of raw imaging data.^10,23^ To complement analyses of clinical and cognitive outcomes, we examined associations between stroke subtype and brain imaging phenotypes, focusing on white matter hyperintensity (WMH) volume.

Analyses of imaging phenotypes were conducted as cross-sectional assessments of post-stroke brain structure. For participants with a history of stroke, MRI scans were obtained after the stroke event; the timing of imaging relative to the stroke varied across individuals. Stroke subtype was defined based on the most recent recorded stroke event occurring before the imaging visit. Stroke-free participants contributed imaging data from their available MRI assessment. We used linear regression models to evaluate differences in WMH volume by stroke subtype among participants with available MRI data, with stroke-free participants serving as the reference group. Models were adjusted for imaging site, baseline systolic and diastolic blood pressure, age at scan, age at scan squared, sex, age-by-sex interaction, ethnicity, and mean relative head motion from resting-state and task-based fMRI. To account for potential nonlinear and interaction effects, we additionally included age at scan squared and an age-by-sex interaction term, consistent with prior UK Biobank imaging analyses.^24^

### Association between stroke type and mortality

We conducted two complementary analyses to examine the association between stroke and mortality, which differed in their study populations and time origins.

### Post-stroke survival analysis

Among participants who experienced a stroke, we estimated post-stroke survival separately for each stroke subtype. Follow-up time was defined as the interval from the index stroke date to death or censoring. Cox proportional hazards models were fit with time since stroke as the time scale and stroke subtype as the stratifying variable (allowing subtype-specific baseline hazards), adjusting for age, sex, ethnicity, systolic and diastolic blood pressure, and body mass index (BMI). The fitted models were used to derive covariate-adjusted survival curves for each stroke subtype. To be included in the survival curve analysis, participants were required to have a valid index stroke date and a non-missing, non-negative follow-up interval (i.e., a death or censoring date occurring after the index stroke); individuals missing these fields or with inconsistent dates were excluded, resulting in a smaller analytic sample for the survival curves than the overall analytic stroke subcohort. As highlighted in Supplementary Table 7, proportional hazards assumptions were evaluated using Schoenfeld residuals; some evidence of non-proportionality was observed for age, sex, and diastolic blood pressure, so results are interpreted as average effects over follow-up.

### Stroke as a time-varying exposure and mortality risk

In a second analysis, we evaluated the association between stroke occurrence and subsequent all-cause mortality in the full UK Biobank cohort, including both stroke-free participants and those who experienced a stroke during follow-up. Stroke status was modeled as a time-varying exposure, with individuals contributing stroke-free person-time before their first stroke and post-stroke person-time thereafter. Follow-up began at the baseline UK Biobank assessment (instance 0) and continued until death or censoring. Separate Cox proportional hazards models were fit for each stroke subtype, comparing participants with that subtype to stroke-free participants. Models were adjusted for age, sex, ethnicity, systolic and diastolic blood pressure, and body mass index (BMI) measured at baseline, with continuous covariates standardized to one standard deviation to facilitate interpretability. Hazard ratios from these models represent the relative hazard of mortality after stroke, compared with stroke-free person-time. Together with the post-stroke survival analysis, this approach allows complementary assessment of long-term prognosis among stroke survivors and the population-level increase in mortality hazard associated with stroke occurrence.

### Standard Protocol Approvals, Registrations, and Patient Consents

This study used deidentified data from the UK Biobank and was conducted under UK Biobank application no. 52887. The secondary analysis was approved by the Harvard Institutional Review Board, Harvard University, under protocol no. IRB16-2145. All UK Biobank participants provided written informed consent for participation and for use of their deidentified health-related data for approved research. Because the present study involved secondary analysis of deidentified data, no additional participant consent was required. No experiments involving live vertebrates or higher invertebrates were performed, and no recognizable participant photographs, videos, or other identifying information are included. This observational study was not a clinical trial; therefore, clinical trial registration and submission of a trial protocol or statistical analysis plan are not applicable.

## Results

### Cohort demographics

Of UKB participants who had a stroke during surveillance (n=14,806), we found that 69.24% of strokes were ischemic, 9.51% subarachnoid, 9.78% ONIH, and 11.5% ICH (Table 1). We examined the baseline clinical and demographic factors associated with each subtype (Table 1). Ischemic, ONIH, and intracerebral strokes occurred more frequently among men, whereas subarachnoid hemorrhage occurred more frequently among women. There is a similar distribution of other demographic factors across stroke types.

**Table 1.** Baseline demographic and clinical characteristics of the analytic stroke cohort by stroke subtype. Values are mean or percentage, as indicated. Cardiovascular PRS (polygenic risk score) values are standardized to mean 0, SD 1, from Field ID 26223 Standard PRS for cardiovascular disease (CVD).

|  | **Ischemic** | **Intracerebral** | **Subarachnoid** | **Other Nontraumatic Intracranial Hemorrhage** |
| --- | --- | --- | --- | --- |
| **N** | 10,251 | 1,699 | 1,408 | 1,448 |
| **Percent of stroke cohort (%)** | 69 | 11.5 | 9.51 | 9.78 |
| **Sex (Male, %)** | 60.6 | 54.6 | 36.7 | 62.6 |
| **Mean Age at Stroke** | 68.5 ± 9.01 | 67.7 ± 10.2 | 56.8 ± 13.5 | 66.4 ± 13.1 |
| **Deceased by the end of follow-up (%)** | 23.6 | 41.4 | 25.4 | 34.9 |
| **European ancestry (%)** | 95 | 94.6 | 94.5 | 94.7 |
| **Non-European ancestry (%)** | 4.9 | 5.4 | 5.5 | 5.3 |
| **Mean BMI (kg/m<sup>2</sup>)** | 28.5 | 27.6 | 27.2 | 27.7 |
| **Diastolic Blood Pressure (mmHg)** | 83.8 | 83.8 | 82.5 | 82.8 |
| **Systolic Blood Pressure (mmHg)** | 145 | 143 | 140 | 142 |
| **LDL (mmol/L)** | 1.61 | 1.64 | 1.68 | 1.56 |
| **HDL (mmol/L)** | 1.23 | 1.26 | 1.33 | 1.25 |
| **Rigorous Physical Activity per Week (days)** | 1.72 | 1.73 | 1.76 | 1.80 |
| **Enhanced Cardiovascular PRS** | 0.171 | -0.05 | -0.07 | -0.13 |

We evaluated post-stroke outcomes across survival, dementia, cognition, and neuroimaging domains. A summary of findings across these complementary dimensions of neurological burden is shown in Figure 2.

**Figure 2.**
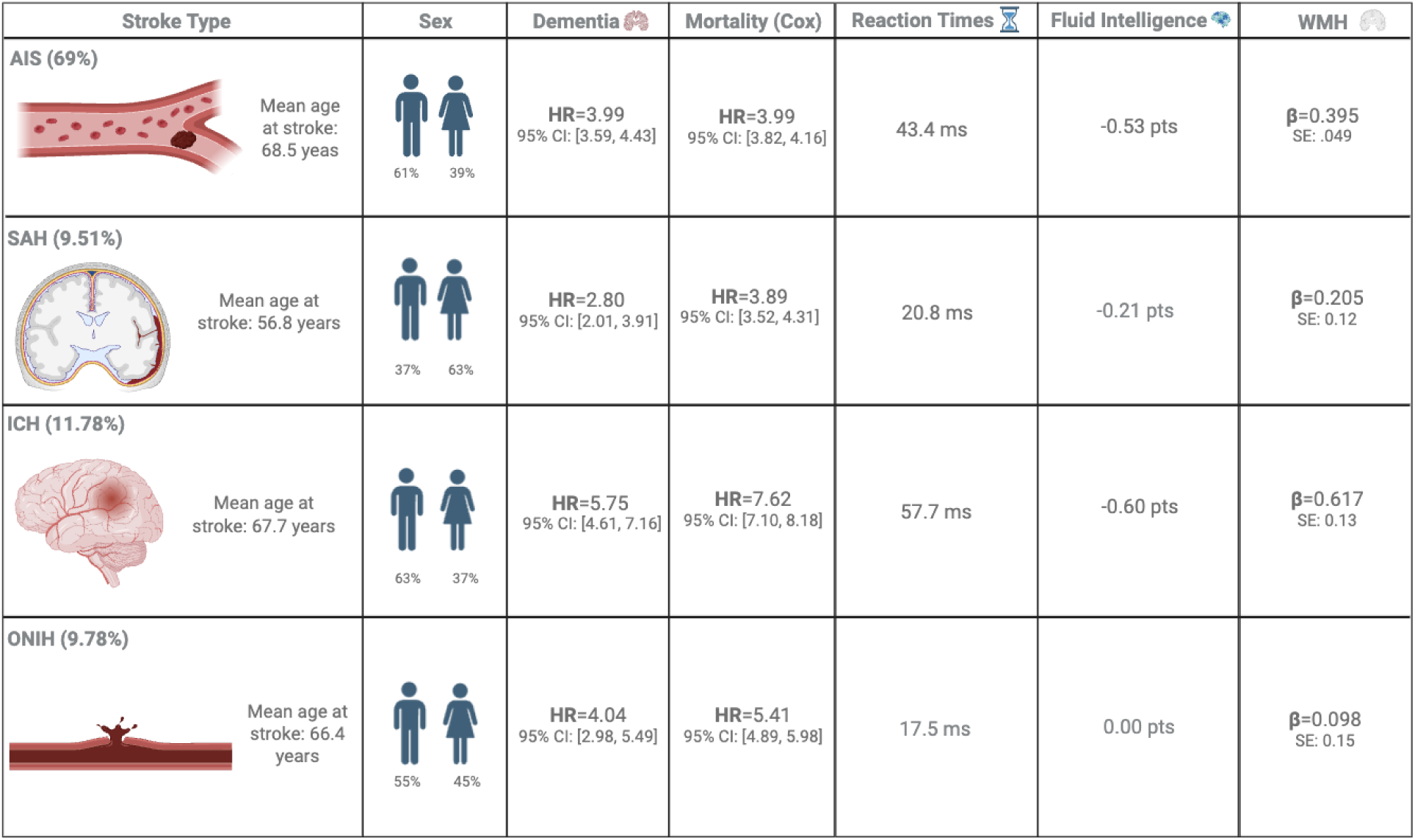
Multidimensional characterization of long-term neurological outcomes following stroke. Stroke was associated with adverse outcomes across survival, neurodegenerative, cognitive, and neuroimaging domains. Estimates summarize associations with mortality, dementia, cognitive performance, and white matter hyperintensity (WMH) burden relative to individuals without stroke. Together, these findings illustrate that post-stroke neurological burden extends across multiple dimensions of brain health and cannot be fully captured by any single outcome measure.

**Figure 3.**
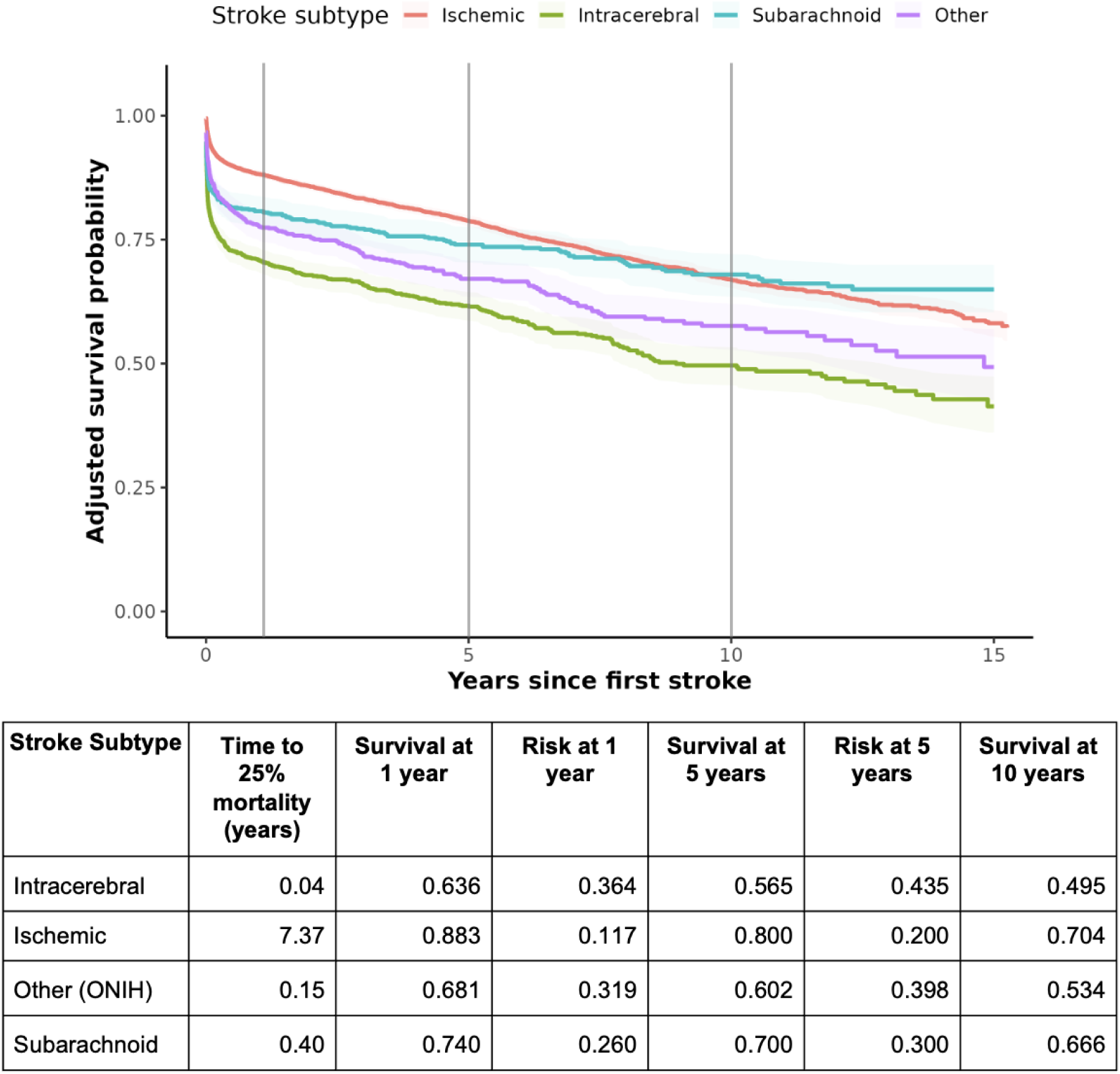
Covariate-adjusted post-stroke survival by stroke subtype. Curves show model-estimated survival probability over 15 years after index stroke for ischemic stroke, intracerebral hemorrhage, subarachnoid hemorrhage, and other nontraumatic intracranial hemorrhage (ONIH). Survival curves were derived from Cox proportional hazards models adjusted for age, sex, ethnicity, systolic blood pressure, diastolic blood pressure, and body mass index. Shaded bands represent 95% confidence intervals. Gray vertical lines mark 1, 5, and 10 years after stroke. The accompanying table summarizes model-estimated survival probabilities at 1, 5, and 10 years, corresponding absolute mortality risk calculated as 1 − S(t), and time to 25% mortality for each stroke subtype.

### Dementia risk following stroke

In a multivariable Cox proportional hazards model including 466,649 participants and 6,626 incident dementia events, all stroke subtypes were associated with a significantly higher hazard of developing dementia compared with stroke-free individuals. Intracerebral hemorrhage was associated with the highest risk of dementia (hazard ratio [HR] = 5.75, 95% CI 4.61–7.16), followed by other nontraumatic intracranial hemorrhage (HR = 4.04, 95% CI 2.98–5.49) and ischemic stroke (HR = 3.99, 95% CI 3.59–4.43). Subarachnoid hemorrhage was associated with a lower, though still significantly elevated, dementia risk (HR = 2.80, 95% CI 2.01–3.91) (Supplementary Table 11).

Among individuals with stroke who developed dementia during follow-up, the median time from stroke to dementia onset was 4.4 years overall, with earlier onset observed after ischemic stroke and intracerebral hemorrhage and later onset after subarachnoid hemorrhage (Supplementary Table 10). Formal proportional hazards testing for the dementia models is shown in Supplementary Table 9.

### Post-stroke dementia incidence accounting for competing mortality

To better characterize dementia risk among stroke survivors, we performed a Fine-Gray competing risks analyses treating death prior to dementia as a competing event.^20^

Among individuals with incident stroke included in the competing risks cohort (n = 12,801), 493 developed dementia during follow-up, while 2,887 died prior to dementia, indicating substantial competing mortality. These findings align with the marked heterogeneity in post-stroke survival observed across stroke subtypes. The Fine–Gray competing-risk cohort was smaller than the descriptive time-to-dementia cohort because it required valid index stroke, dementia, death, and censoring dates with non-negative follow-up time; Supplementary Table 10 summarizes time to dementia among the broader set of stroke participants who developed dementia during follow-up.

Cumulative incidence analyses demonstrated that dementia incidence following stroke was modest relative to mortality. At 5 years post-stroke, the cumulative incidence of dementia ranged from approximately 1% for subarachnoid hemorrhage to 3–4% for ischemic and intracerebral stroke, whereas the cumulative incidence of death ranged from approximately 18% to over 30% across subtypes (Figure 4). This divergence persisted over longer follow-up, with mortality substantially exceeding dementia incidence in all stroke groups.

**Figure 4.**
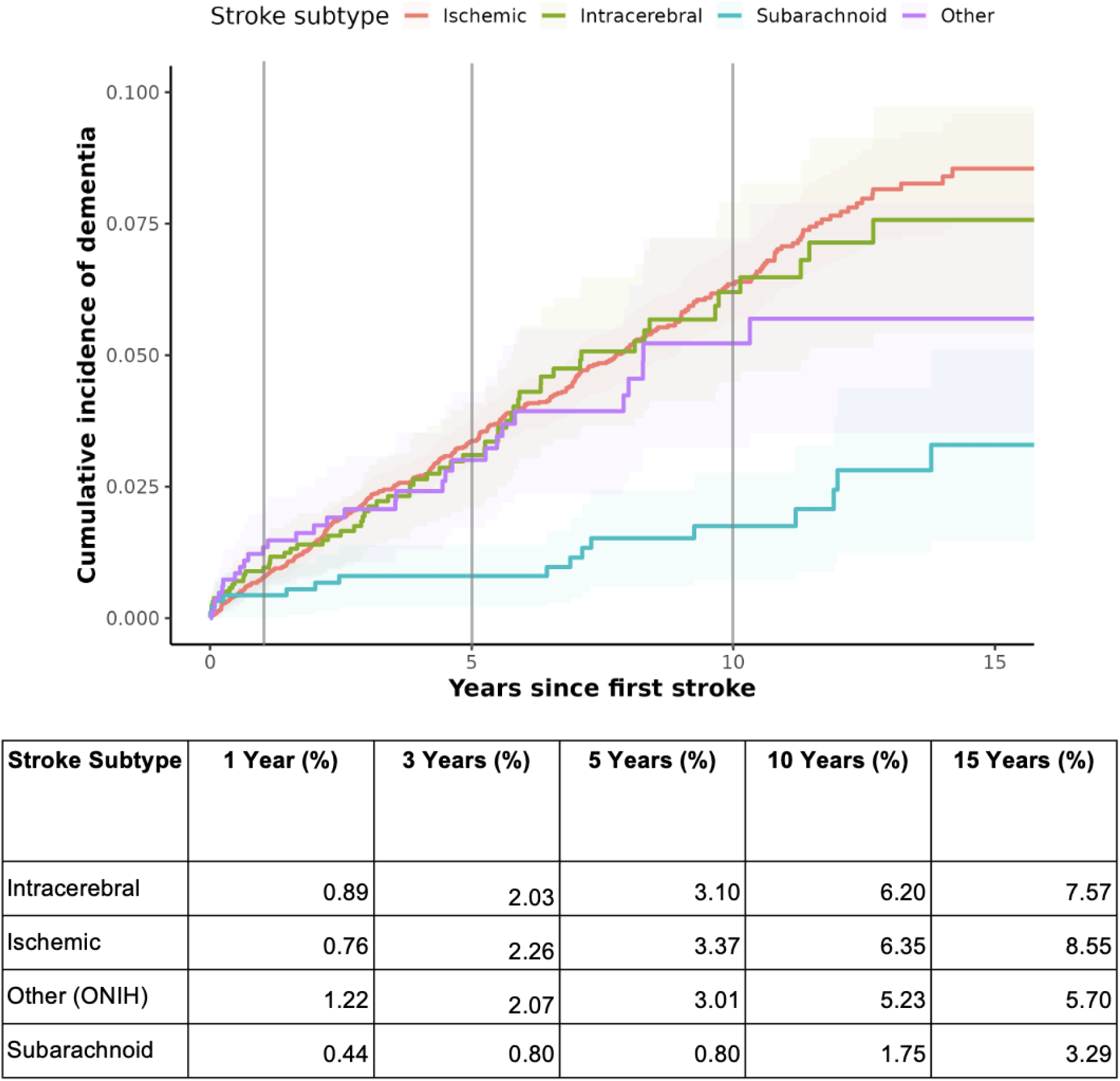
Cumulative incidence of all-cause dementia following index stroke by first stroke subtype. Cumulative incidence functions (CIFs) are shown for ischemic stroke, intracerebral hemorrhage, subarachnoid hemorrhage, and other nontraumatic intracranial hemorrhage. Time is measured from the index stroke event. Death prior to dementia was treated as a competing event. The accompanying table summarizes estimated cumulative incidence of dementia at 1, 3, 5, 10, and 15 years following stroke.

In competing risks regression models, subarachnoid hemorrhage was associated with a significantly lower cumulative incidence of dementia compared with ischemic stroke (subdistribution hazard ratio [sHR] 0.47, 95% CI 0.28–0.77, p = 0.003), whereas intracerebral hemorrhage and other nontraumatic intracranial hemorrhage were not significantly different from ischemic stroke. Older age at stroke was strongly associated with increased dementia incidence (sHR 1.06 per year, p < 0.001), and male sex was associated with a higher risk (sHR 1.28, p = 0.009). African ethnicity was also associated with higher dementia incidence, while other ethnicity categories were not statistically significant (Figure 6).

**Figure 5:**
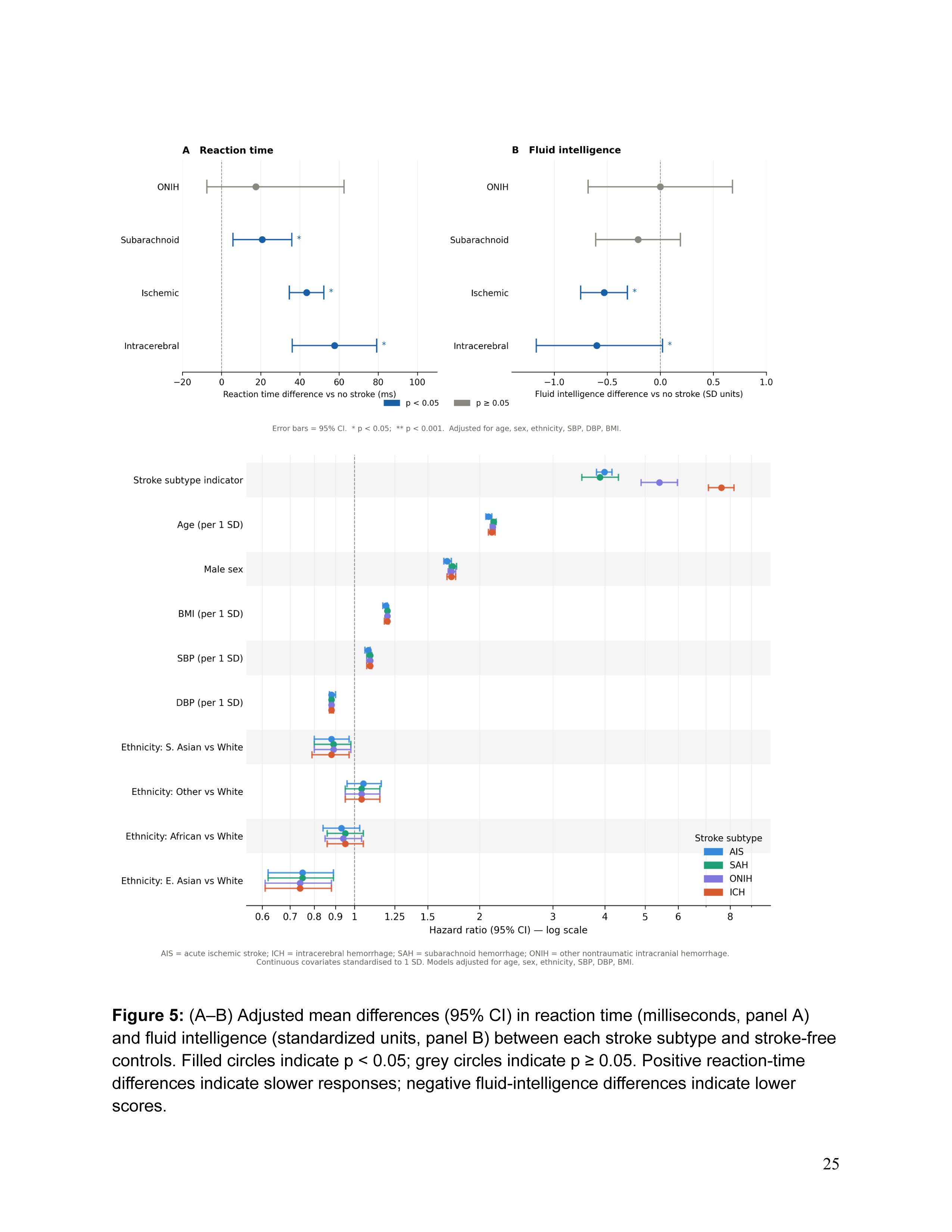
(A–B) Adjusted mean differences (95% CI) in reaction time (milliseconds, panel A) and fluid intelligence (standardized units, panel B) between each stroke subtype and stroke-free controls. Filled circles indicate p < 0.05; grey circles indicate p ≥ 0.05. Positive reaction-time differences indicate slower responses; negative fluid-intelligence differences indicate lower scores.

**Figure 6:**
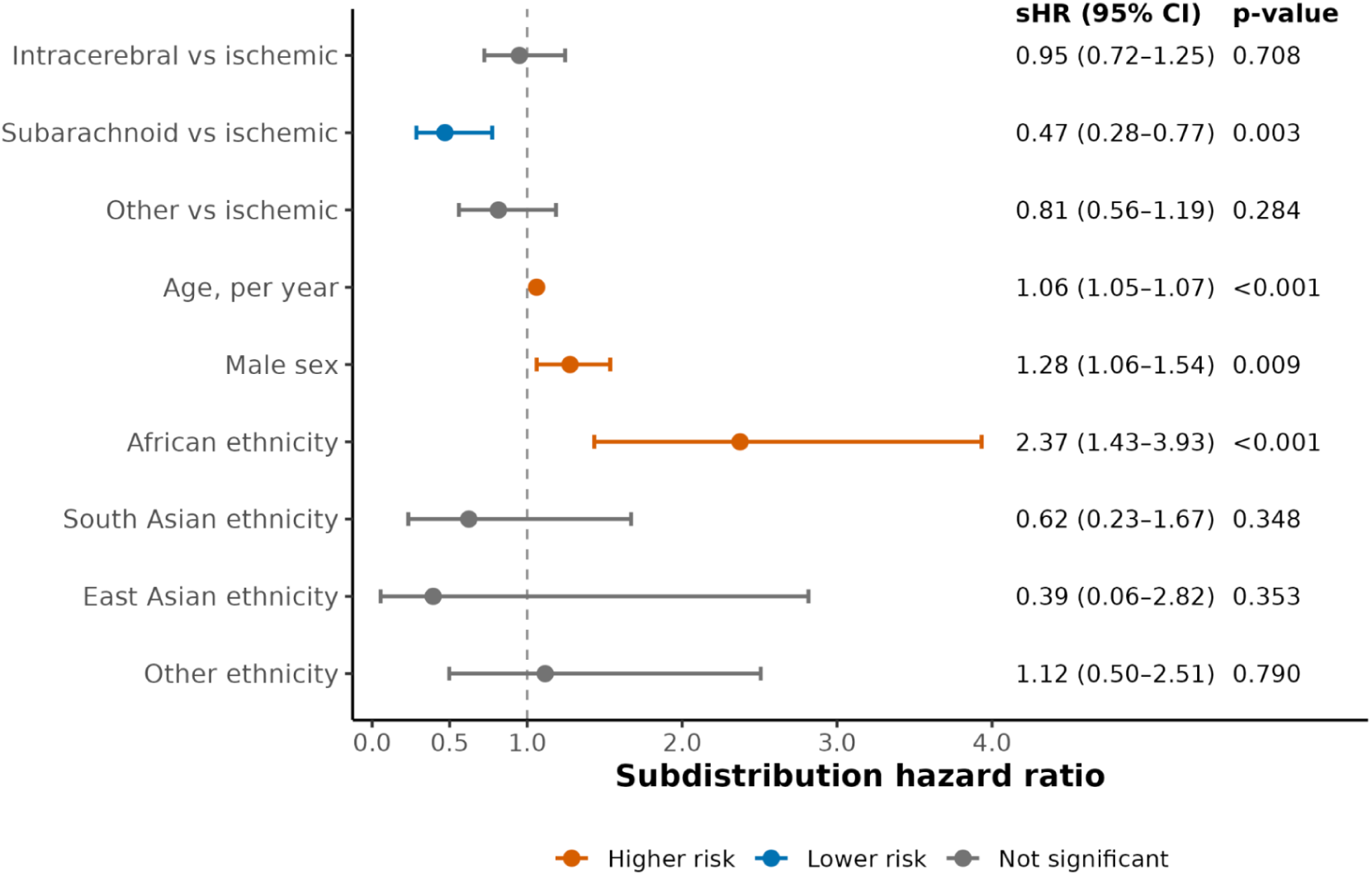
Subdistribution hazard ratios (sHRs) for incident dementia following stroke from Fine–Gray competing risks models accounting for death as a competing event.

These findings should be interpreted in the context of substantial competing mortality, particularly among hemorrhagic stroke subtypes, where elevated early mortality may limit the observed cumulative incidence of dementia. Results were consistent in sensitivity analyses adjusting for baseline cardiovascular risk factors and in cause-specific Cox models.

### Post-stroke participants exhibit significantly longer reaction times

Figure 5 shows adjusted cognitive differences by stroke subtype relative to stroke-free controls. Overall, the reaction times increased significantly for participants post-stroke, with considerable variability between stroke types. The timing of cognitive assessment after stroke varied across participants and by stroke subtype, as summarized in Supplementary Table 12. All cognitive comparisons reflect cross-sectional post-stroke performance rather than within-individual cognitive change over time.

Relative to the no-stroke group, post-stroke participants exhibited significantly longer reaction times. Larger effect sizes were observed in certain stroke groups, including intracerebral and ischemic stroke, while smaller increases were observed in others (Figure 5; Supplementary Tables 2–5).

### Post-stroke participants exhibit lower cognitive reasoning

There were significant differences in reasoning scores between stroke-exposed and stroke-free participants (Figure 5B). Post-stroke participants demonstrated lower cognitive scores relative to stroke-free individuals. Reductions were more pronounced in certain groups, including intracerebral and ischemic stroke, while other groups showed no significant differences. Individuals with intracerebral hemorrhage showed a mean difference of −0.606 points (95% CI, −1.14 to −0.07). Similarly, individuals with ischemic stroke had a mean difference of −0.541 points (95% CI, −0.75 to −0.33). In contrast, some groups did not show significant differences. Cognitive scores among individuals with subarachnoid hemorrhage did not differ significantly from those of stroke-free participants.

In addition to reasoning and reaction time, we evaluated other cognitive outcomes, including symbol digit substitution, matrix pattern completion, numeric trail making, and numeric memory, shown in Supplementary Tables 2–5.

### White matter hyperintensity burden following stroke

In multivariable linear regression models with normalized white matter hyperintensity (WMH) volume as the outcome, stroke exposure was associated with higher post-stroke WMH burden. Ischemic stroke was associated with a 0.40-SD higher WMH volume (β = 0.395; 95% CI, 0.30–0.49), while intracerebral hemorrhage was associated with a 0.62-SD higher WMH volume (β = 0.617; 95% CI, 0.36–0.87). Associations were not statistically significant for SAH or ONIH.

Higher baseline blood pressure was also independently associated with greater WMH burden. Each standard deviation increase in mean diastolic blood pressure was associated with a 0.06-SD increase in WMH volume (β = 0.058; 95% CI, 0.04–0.07), and each standard deviation increase in mean systolic blood pressure was associated with a 0.04-SD increase in WMH volume (β = 0.037; 95% CI, 0.02–0.05).

Age at scan demonstrated a strong nonlinear association with WMH volume, with significant linear and quadratic age terms (both p < 0.001). Female sex was associated with a modestly lower WMH burden than in males (β = −0.021; 95% CI, −0.04 to −0.001), whereas the age-by-sex interaction was not significant. The fully adjusted model explained approximately 18% of the variance in WMH volume (R² = 0.18). Full regression results are presented in Supplementary Table 6.

### Mortality following stroke

We observed substantial heterogeneity in post-stroke survival outcomes (Figure 3). Early mortality varied substantially across individuals and stroke characteristics. Among individuals aged 45 and older, 25% mortality occurred within 0.03 years (∼11 days) for ICH (95% CI: 0.022–0.063), 0.14 years (∼52 days) for ONIH (95% CI: 0.06–0.29), 0.39 years (∼142 days) for SAH (95% CI: 0.07–2.37), and 7.36 years for AIS (95% CI: 6.83–8.06).

Stroke was associated with a substantially elevated mortality hazard compared with stroke-free person-time. In time-varying Cox models, ischemic stroke was associated with a nearly four-fold increase in post-stroke mortality hazard compared with stroke-free person-time (HR 3.99, 95% CI 3.82–4.16). The magnitude of mortality risk varied across groups. Intracerebral hemorrhage was associated with the highest mortality hazard (HR 7.62, 95% CI 7.10–8.18), followed by other nontraumatic intracranial hemorrhage (HR 5.41, 95% CI 4.89–5.98) and subarachnoid hemorrhage (HR 3.89, 95% CI 3.52–4.31). Overall, these findings indicate marked heterogeneity in post-stroke mortality hazard across stroke subtypes, with intracerebral hemorrhage conferring the greatest risk of death.

## Discussion

In this large population-based cohort, we evaluated long-term neurological outcomes following stroke across survival, dementia, cognitive performance, and neuroimaging phenotypes. Although each of these outcomes has been linked to stroke, they are often examined in isolation.^4–11,14^ By evaluating them within a single population, we found that the long-term neurological burden of stroke spans multiple dimensions of brain health and cannot be fully captured by any single endpoint. Mortality, dementia, cognitive performance, and white matter hyperintensity burden each reflected distinct aspects of post-stroke prognosis, highlighting the value of integrated approaches to understanding long-term outcomes after stroke.

A central observation was the divergence between mortality and neurological outcomes. Mortality risk following stroke was substantial, particularly in the early period after hemorrhagic events, such that death frequently preceded the development of dementia. Consequently, estimates of post-stroke dementia burden depended strongly on whether mortality was incorporated into the analytic framework. Unlike cause-specific hazard models, which condition on survival, competing-risk approaches account for the fact that death may preclude the observation of dementia. Although stroke was associated with elevated dementia risk, the cumulative incidence of dementia remained modest relative to mortality across all stroke groups, indicating that competing mortality substantially shapes the observed burden of post-stroke neurodegenerative disease. These findings emphasize that long-term neurological outcomes cannot be interpreted independently of survival.

Among individuals who survived the acute post-stroke period, we observed consistent evidence of persistent neurological impairment across cognitive and neuroimaging domains. Cognitive performance differed across domains, with impairments in processing speed and reasoning observed at post-stroke assessments occurring several years after the index event. These analyses were cross-sectional and therefore reflect residual or persistent deficits rather than within-individual cognitive decline.^9, 25,26^ This distinction is important: post-stroke cognitive performance captures functional outcomes at a given time point, whereas dementia represents a progressive clinical syndrome that may emerge over extended follow-up. The dissociation between cognitive performance and dementia incidence observed in this cohort suggests that these outcomes represent related, but non-equivalent, manifestations of post-stroke neurological injury.

For example, we observed significant decreases in reasoning scores among post-stroke participants, with larger effects in certain groups. Reasoning scores reflect an individual’s ability to solve problems that require logical reasoning, independent of acquired knowledge or educational attainment, and are closely linked to memory and attentional processes. Reaction time and fluid intelligence capture distinct cognitive domains: reaction time reflects processing speed and sensorimotor integration, whereas fluid intelligence emphasizes reasoning and problem-solving. Differences between these outcomes may therefore reflect domain-specific vulnerability rather than global cognitive impairment.^27^

Importantly, cognitive analyses in this study were based on the closest available post-stroke assessment. As such, observed differences reflect post-stroke cognitive performance rather than within-individual cognitive trajectories. Performance on specific tasks may also be influenced by residual motor, language, or visual deficits following stroke, potentially contributing to variation across groups and to selection bias favoring individuals able to complete testing.^28^ These considerations are particularly relevant when interpreting reaction time and executive-function tasks in populations with variable neurological deficits. More broadly, cognitive and imaging analyses were restricted to participants who survived long enough and were healthy enough to undergo assessment, raising the possibility that the burden of post-stroke neurological impairment is underestimated in this cohort.

In addition to cognitive differences, we observed heterogeneity in the risk of incident dementia following stroke. Intracerebral hemorrhage was associated with the highest subsequent dementia risk, followed by ONIH and ischemic stroke, whereas subarachnoid hemorrhage exhibited comparatively lower long-term dementia risk among survivors.^29^ These findings are consistent with prior work suggesting that post-stroke dementia reflects a combination of acute injury severity, secondary neurodegenerative processes, and underlying cerebrovascular vulnerability.^5–7^ Hemorrhagic strokes, particularly ICH, may confer elevated dementia risk through greater parenchymal injury, disruption of subcortical networks, and a higher burden of pre-existing small vessel disease.^10,30^

Neuroimaging findings further support the presence of persistent neurological injury following stroke. White matter hyperintensity burden was elevated among stroke survivors, particularly among individuals with ischemic stroke and intracerebral hemorrhage, consistent with the contribution of chronic small-vessel disease and secondary white matter injury.^10,11^ Together with the cognitive findings, these results suggest that long-term neurological burden extends beyond clinically recognized dementia and may be detectable through both functional and structural measures of brain health.

Several limitations should be considered. Stroke severity measures, including NIHSS scores, lesion volume, hematoma burden, and functional status, were not available in the UK Biobank and may account for some variation in long-term outcomes.^31,32^ The index stroke represented the first recorded stroke within UK Biobank rather than a confirmed first-ever event. Cognitive and neuroimaging analyses were cross-sectional and therefore reflect post-stroke performance and brain structure rather than within-individual trajectories. In addition, cognitive testing and MRI were available only among participants who survived and completed follow-up assessments, which may have led to an underestimation of post-stroke neurological impairment.

Our results support multidimensional approaches to post-stroke monitoring and prognostication while emphasizing the importance of careful interpretation given differences in timing, selection, and outcome measurement across domains. Future studies incorporating measures of stroke severity, lesion burden, and longitudinal cognitive follow-up may provide a more complete understanding of post-stroke neurological trajectories and help guide individualized rehabilitation and surveillance strategies.

Together, these findings suggest that long-term neurological burden after stroke extends beyond any single clinical outcome and is best understood through complementary assessments of survival, cognition, neurodegeneration, and structural brain injury.

## Data Availability

The data used in this study are available from the UK Biobank under application-based access.

https://www.ukbiobank.ac.uk/

## Non-standard Abbreviations and Acronyms

AIS: Acute ischemic stroke
SAH: Subarachnoid hemorrhage
ICH: Intracerebral hemorrhage
ONIH: Other nontraumatic intracranial hemorrhage
UKB: UK Biobank
WMH: White matter hyperintensity
HR: Hazard ratio

## Acknowledgements

We want to acknowledge the UK Biobank for providing us with access to the data they collected. The UK Biobank project number is 52887. Funding sources included the National Institutes of Environmental Health Sciences (NIEHS) R01ES032470, National Institute of Diabetes and Digestive and Kidney Diseases (NIDDK) R01DK137993, National Institutes of Environmental Health Sciences (NIEHS) U24ES036819, and Advanced Research Projects Agency for Health ARPA-H D24AC00345. The funders had no role in the research.

